# A Multi-Domain EEG Framework Integrating Spectral, Functional Connectivity, and Topological Features for Subject-Level ADHD Classification

**DOI:** 10.64898/2026.10.08.26365043

**Authors:** Faezeh Rohani, Sayediman Mostafavi, Payam Ebrahimifakhar, Reza Zomorrodi, Andreas Mueller

## Abstract

**Background:** Attention-Deficit/Hyperactivity Disorder (ADHD) is a common neurodevelopmental disorder characterized by persistent symptoms of inattention, hyperactivity, and impulsivity that significantly affect cognitive, academic, and social functioning. Although electroencephalography (EEG) has emerged as a promising non-invasive tool for identifying objective neurophysiological biomarkers of ADHD because of its high temporal resolution and cost-effectiveness, many existing machine learning studies rely on single-domain EEG features, which may not adequately capture the complex functional organization of the brain.

**Methods:** This study proposes a subject-level EEG classification framework that integrates three complementary feature domains: spectral characteristics, weighted Phase Lag Index (wPLI)-based functional connectivity, and topological descriptors derived from persistent homology. EEG recordings from 121 subjects were preprocessed and segmented into overlapping windows for feature extraction. Window-level features were aggregated to generate subject-level representations. Recursive Feature Elimination (RFE) was employed to select the 110 most informative features from an initial set of 1,043 descriptors. Five machine learning classifiers, including Support Vector Machine (SVM), Random Forest, XGBoost, CatBoost, and Multi-Layer Perceptron (MLP), were evaluated using stratified five-fold cross-validation. An ablation study was further conducted to assess the contribution of each feature domain. Model interpretability was investigated using SHapley Additive exPlanations (SHAP) and Local Interpretable Model-agnostic Explanations (LIME), two widely adopted Explainable Artificial Intelligence techniques for interpreting machine learning models.

**Results:** Among the evaluated classifiers, the SVM achieved the highest classification performance, yielding an accuracy of 80.17%, an F1-score of 80.95%, a balanced accuracy of 80.14%, a receiver operating characteristic area under the curve (ROC-AUC) of 86.72%, and a precision–recall area under the curve (PR-AUC) of 87.34%. The ablation study demonstrated that combining spectral, functional connectivity, and topological features consistently outperformed individual feature domains, highlighting the complementary nature of these representations. Explainable AI analyses further identified functional connectivity features as the primary contributors to classification performance while confirming the additional value of spectral and topological descriptors.

**Conclusions:** The proposed framework demonstrates that integrating spectral analysis, functional connectivity, and topological data analysis provides an effective and interpretable approach for subject-level ADHD classification from EEG recordings. The combination of recursive feature selection, multi-domain feature fusion, ablation analysis, and Explainable Artificial Intelligence establishes a transparent and reproducible pipeline that may facilitate the development of reliable EEG-based clinical decision-support systems for ADHD.

## 1. Introduction

Attention-Deficit/Hyperactivity Disorder (ADHD) is one of the most prevalent neurodevelopmental disorders, affecting approximately 5–7% of children worldwide and frequently persisting into adolescence and adulthood [1–3]. The disorder is characterized by persistent patterns of inattention, hyperactivity, and impulsivity that negatively affect academic achievement, occupational performance, and social functioning [1,3]. Although the diagnosis of ADHD is primarily based on clinical interviews, behavioral observations, and standardized diagnostic criteria, these approaches remain largely subjective and may be influenced by clinician experience and patient behavior during assessment [1]. Consequently, considerable effort has been devoted to identifying objective neurophysiological biomarkers capable of supporting clinical diagnosis and improving diagnostic reliability [4].

Electroencephalography (EEG) has emerged as one of the most promising non-invasive techniques for investigating the neural mechanisms underlying ADHD because of its excellent temporal resolution, relatively low acquisition cost, portability, and widespread clinical availability [5–7]. Numerous studies have demonstrated that individuals with ADHD exhibit abnormalities in neural oscillatory activity, particularly within the delta, theta, alpha, beta, and gamma frequency bands, reflecting alterations in cortical maturation and functional brain organization [5,7,8]. Traditional EEG biomarkers, including the theta-to-beta ratio, have shown potential for ADHD assessment; however, their diagnostic performance remains inconsistent across different datasets and populations, indicating that single-domain EEG features may not sufficiently characterize the complex neural dynamics associated with ADHD [8,9].

To overcome these limitations, machine learning techniques have been increasingly applied to EEG analysis for automated ADHD classification [10–12]. Supervised learning algorithms, including Support Vector Machines (SVM), Random Forests, Gradient Boosting methods, Artificial Neural Networks, and Deep Learning models, have demonstrated encouraging performance in distinguishing individuals with ADHD from healthy controls [11–13]. Despite these advances, many existing studies rely primarily on spectral or temporal EEG features while overlooking complementary information contained within functional brain connectivity and higher-order network organization [11,14]. Furthermore, a substantial proportion of published studies perform classification at the EEG window level, allowing multiple windows from the same participant to contribute independently to model evaluation. This approach may inflate classification performance because windows originating from the same subject are not statistically independent [15,16].

Functional connectivity analysis provides an alternative representation of brain activity by quantifying interactions between spatially distributed cortical regions rather than examining individual EEG channels independently [17,18]. Among the available connectivity measures, the weighted Phase Lag Index (wPLI) has gained considerable attention because it reduces the influence of volume conduction and common-source artifacts while providing reliable estimates of phase synchronization between brain regions [19]. However, functional connectivity alone does not fully describe the global structural organization of brain networks.

Topological Data Analysis (TDA), particularly persistent homology, has recently emerged as a powerful mathematical framework for characterizing complex network organization across multiple spatial scales [20–22]. Unlike conventional graph-theoretical approaches that require arbitrary threshold selection, persistent homology captures stable topological structures directly from weighted connectivity networks, enabling robust characterization of higher-order brain organization [20,23]. Although TDA has shown promising results in several neurological and psychiatric disorders, its application to EEG-based ADHD classification remains relatively limited, particularly in combination with conventional spectral and functional connectivity features.

In addition to achieving high classification accuracy, the interpretability of machine learning models has become increasingly important in biomedical applications. Black-box models may achieve strong predictive performance while providing limited insight into the physiological factors driving their predictions, reducing their clinical interpretability and trustworthiness [24]. Explainable Artificial Intelligence (XAI) techniques such as SHapley Additive exPlanations (SHAP) and Local Interpretable Model-agnostic Explanations (LIME) provide complementary global and local explanations of machine learning predictions, enabling the identification of influential EEG biomarkers and improving the transparency of diagnostic models [25–27].

In this study, we propose a comprehensive subject-level EEG classification framework that integrates spectral features, weighted Phase Lag Index (wPLI)-based functional connectivity, and topological descriptors derived from persistent homology. Following EEG preprocessing and window segmentation, window-level features are aggregated to construct subject-level representations. Recursive Feature Elimination (RFE) is subsequently employed to identify the most informative features prior to classification using multiple machine learning algorithms. The proposed framework is further evaluated through an ablation study to quantify the contribution of each feature domain, while SHAP and LIME are used to provide both global and local explanations of the final classification model. The overall workflow of the proposed framework is illustrated in Figure 1. By combining complementary EEG feature representations, rigorous feature selection, comprehensive model evaluation, and Explainable Artificial Intelligence, this study aims to provide a robust, interpretable, and reproducible framework for EEG-based ADHD classification that may support the development of objective clinical decision-support systems.

**Figure 1.**
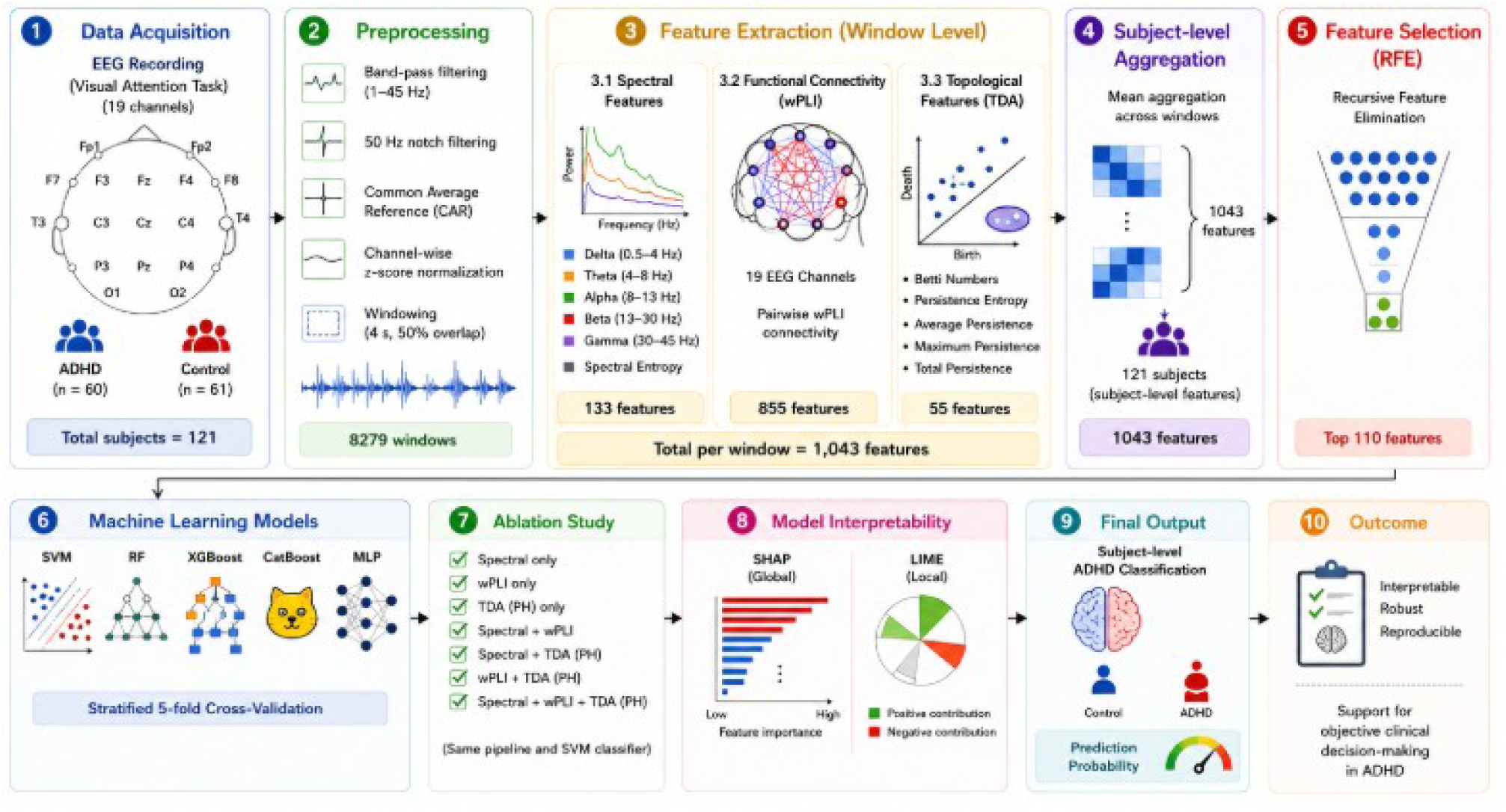
Overview of the proposed explainable subject-level EEG classification framework for ADHD detection. EEG recordings from 121 participants (61 ADHD and 60 healthy controls) were preprocessed using 1–45 Hz band-pass filtering, 50 Hz notch filtering, Common Average Reference (CAR), channel-wise z-score normalization, and segmentation into 4-second windows with 50% overlap. Three complementary feature domains were extracted from each EEG window: spectral features (133), weighted Phase Lag Index (wPLI)- based functional connectivity features (855), and persistent homology-based topological descriptors (55), yielding a total of 1,043 features. Window-level features were aggregated into subject-level representations, followed by Recursive Feature Elimination (RFE) to select the 110 most informative features. Five machine learning classifiers were evaluated using stratified five-fold cross-validation, while an ablation study quantified the contribution of each feature domain. Finally, SHAP and LIME were employed to provide global and local explanations of the best-performing model, enabling interpretable subject-level ADHD classification.

## 2. Materials and Methods

### 2.1 Dataset

The present study utilized a publicly available EEG dataset for Attention-Deficit/Hyperactivity Disorder (ADHD), accessed through the Kaggle platform and derived from the open-access IEEE DataPort ADHD EEG database [28]. The dataset consists of EEG recordings from 121 children between 7 and 12 years of age, including 61 children diagnosed with ADHD and 60 healthy control participants. ADHD diagnoses were established by experienced psychiatrists according to the Diagnostic and Statistical Manual of Mental Disorders, Fourth Edition (DSM-IV) criteria. The control group consisted of children without a history of psychiatric disorders, epilepsy, or other neurological conditions.

EEG signals were acquired using the international 10–20 electrode placement system with 19 scalp electrodes at a sampling frequency of 128 Hz. The recordings were obtained during a visual attention task, in which participants counted cartoon characters presented on a computer screen while EEG activity was continuously recorded. This experimental paradigm was designed to engage attentional processes relevant to ADHD assessment.

In the present study, all EEG recordings were processed using a unified preprocessing pipeline before feature extraction to ensure consistency across participants. The objective was to perform subject-level binary classification of ADHD and healthy control participants by integrating complementary EEG feature domains, including spectral characteristics, weighted Phase Lag Index (wPLI)-based functional connectivity, and persistent homology-based topological descriptors.

### 2.2 EEG Preprocessing

The raw EEG recordings were preprocessed using a standardized pipeline implemented in Python to improve signal quality and ensure consistency across all subjects. Each EEG window was first band-pass filtered between 1 and 45 Hz using a fourth-order zero-phase Butterworth filter to remove slow baseline drift and high-frequency noise. A 50 Hz notch filter was subsequently applied to suppress power-line interference. The signals were then re-referenced using the Common Average Reference (CAR) method to reduce reference-dependent bias across channels. Finally, channel-wise z-score normalization was performed by subtracting the mean and dividing by the standard deviation of each channel, resulting in zero-mean and unit-variance signals prior to feature extraction.

No automatic or manual artifact rejection, bad-channel interpolation, or epoch rejection procedures were applied. Consequently, all segmented EEG windows were retained for subsequent analysis.

Following preprocessing, each EEG recording was segmented into overlapping temporal windows. Window-based analysis increases the number of available observations while preserving local temporal dynamics, enabling more reliable estimation of spectral, functional connectivity, and topological features [29,30]. All windows were processed independently during feature extraction before being aggregated into subject-level representations.

### 2.3 Window Segmentation

Following preprocessing, each EEG recording was segmented into overlapping temporal windows to facilitate robust feature extraction while preserving the temporal dynamics of the EEG signals. A fixed window length of 4 seconds (512 samples at a sampling frequency of 128 Hz) with 50% overlap (hop size of 256 samples) was employed. This windowing strategy increases the number of observations available for analysis while maintaining sufficient temporal continuity for reliable estimation of spectral, functional connectivity, and topological features.

Each window was treated as an independent sample during the feature extraction stage. Spectral, weighted Phase Lag Index (wPLI)-based functional connectivity, and topological features were extracted separately from every window. To obtain subject-level representations for classification, the feature vectors extracted from all windows belonging to the same subject were averaged, resulting in a single representative feature vector per subject. This aggregation strategy reduces the influence of window-specific fluctuations while preserving the overall neurophysiological characteristics of each participant.

### 2.4 Feature Extraction

To comprehensively characterize EEG activity, three complementary feature domains were extracted from each segmented EEG window: spectral features, functional connectivity features, and topological descriptors derived from persistent homology. These feature domains capture different aspects of brain activity, including local neural oscillations, synchronization between cortical regions, and the global topological organization of functional brain networks. Integrating these complementary representations enables a more comprehensive description of EEG dynamics than relying on a single feature domain alone [31,32].

Feature extraction was performed independently for every EEG window. The resulting window-level feature vectors were subsequently aggregated to generate a single subject-level representation for each participant before feature selection and machine learning classification.

**Table 1.** Summary of the EEG dataset and extracted feature sets.

| Category | Description |
| --- | --- |
| Dataset | Public EEG dataset |
| Number of subjects | 121 |
| ADHD subjects | 61 |
| Healthy controls | 60 |
| EEG channels | 19 (International 10–20 system) |
| Classification task | Binary (ADHD vs. Healthy Control) |
| Analysis level | Subject-level classification |
| Window length | 4 seconds |
| Window overlap | 50% |
| Samples per window | 512 |
| Total EEG windows | 8,279 |
| Spectral features | 133 |
| Functional connectivity (wPLI) features | 855 |
| Persistent homology (TDA) features | 55 |
| Total extracted features | 1,043 |
| Feature selection method | Recursive Feature Elimination (RFE) |
| Selected features | 110 |

#### 2.4.1 Spectral Feature Extraction

Spectral analysis was performed to quantify the oscillatory characteristics of the EEG signals within physiologically meaningful frequency bands. Neural oscillations have been extensively associated with cognitive processing and are known to exhibit abnormal patterns in individuals with ADHD, making spectral analysis one of the most widely adopted approaches for EEG-based ADHD assessment [5,33].

For each segmented EEG window, the power spectral density (PSD) was estimated using Welch’s method [30]. Based on the estimated PSD, relative spectral power was computed for the five conventional EEG frequency bands: delta (1–4 Hz), theta (4–8 Hz), alpha (8–13 Hz), beta (13–30 Hz), and gamma (30–45 Hz). Relative band power provides a normalized representation of oscillatory activity and has been widely used to investigate functional alterations associated with ADHD [5,8].

In addition to relative band power, spectral entropy was calculated for each EEG channel to quantify the complexity and irregularity of the spectral distribution. Spectral entropy provides complementary information regarding signal organization and has been shown to reflect changes in neural dynamics associated with neurological and neurodevelopmental disorders [34,35].

The extracted spectral descriptors therefore consisted of channel-wise relative band powers, spectral entropy, and peak frequency, providing a comprehensive representation of local oscillatory brain activity.

#### 2.4.2 Functional Connectivity Analysis

Functional connectivity was quantified using the Weighted Phase Lag Index (wPLI), which estimates phase synchronization between EEG channels while reducing the influence of volume conduction and common-source artifacts [19]. For each subject, wPLI was computed from the preprocessed EEG windows using the spectral_connectivity_epochs function provided by the MNE-Connectivity library [36]. Spectral connectivity was estimated using the multitaper method, and connectivity values were averaged within each frequency band (delta: 2–4 Hz, theta: 4–8 Hz, alpha: 8–13 Hz, beta: 13–30 Hz, and gamma: 30–45 Hz). For functional connectivity estimation, the delta band was defined as 2–4 Hz to improve the stability of phase-based synchronization estimates near the lower frequency limit of the preprocessed EEG signals. This definition differs slightly from the spectral analysis, where the conventional 1–4 Hz delta band was used for power estimation.

For each subject, all preprocessed EEG windows were treated as epochs for connectivity estimation, resulting in one 19 × 19 symmetric connectivity matrix for each frequency band. The upper triangular elements of each matrix, excluding the diagonal, were retained as unique connectivity values, resulting in 171 connectivity features per frequency band and a total of 855 functional connectivity features per subject.

The extracted functional connectivity features constitute the second feature domain of the proposed framework and provide complementary information to the spectral descriptors by characterizing communication patterns among distributed cortical regions.

#### 2.4.3 Topological Feature Extraction Using Persistent Homology

While functional connectivity characterizes pairwise interactions between brain regions, it does not fully describe the global organization of functional brain networks. To capture higher-order topological structures, Topological Data Analysis (TDA) was performed using persistent homology, a mathematical framework that identifies topological features that remain stable across multiple connectivity thresholds without requiring the selection of a single network threshold [20,22,23].

For each subject and frequency band, the corresponding wPLI connectivity matrix was converted into a distance matrix using 1−wPLI, where stronger functional connections correspond to shorter distances. The resulting distance matrix was used to construct a Vietoris–Rips complex, and persistent homology was computed up to dimension 2 with a maximum edge length of 1.0. Persistence diagrams were subsequently generated to characterize the evolution of topological structures across the filtration process.

From the persistence diagrams, quantitative descriptors were extracted from both 0-dimensional (H₀) and 1-dimensional (H₁) homology. For H₀, five descriptors were computed: maximum lifetime, mean lifetime, median lifetime, total persistence, and persistence entropy. For H₁, six descriptors were extracted, including the number of persistent loops together with the maximum lifetime, mean lifetime, median lifetime, total persistence, and persistence entropy. Consequently, 11 topological descriptors were obtained for each frequency band, yielding a total of 55 topological features per subject across the five EEG frequency bands.

### 2.5 Subject-Level Feature Aggregation

Feature extraction was initially performed independently for each segmented EEG window, resulting in a large number of window-level feature vectors for every participant. Since multiple windows originating from the same subject are highly correlated, directly using these windows for classification may lead to information leakage and overly optimistic estimates of classification performance. To address this issue, a subject-level aggregation strategy was employed to construct a single representative feature vector for each participant [15,16].

For every extracted feature, statistical aggregation was performed across all windows belonging to the same subject. Specifically, the mean value of each feature was calculated across the available windows, producing a stable representation of the subject’s overall EEG characteristics while reducing the influence of transient fluctuations and noise. This aggregation procedure transformed the window-level feature space into a subject-level dataset suitable for machine learning classification.

Following aggregation, each participant was represented by a single feature vector containing the combined spectral, functional connectivity, and topological descriptors. Consequently, the final dataset consisted of 121 subject-level feature vectors, each associated with a single class label corresponding to either the ADHD or healthy control group.

Subject-level aggregation improves the clinical relevance of the proposed framework by ensuring that each participant contributes only one observation during model training and evaluation. This strategy reduces the risk of data leakage between training and testing sets and provides a more realistic estimate of the model’s ability to generalize to previously unseen subjects.

### 2.6 Feature Selection

The integration of spectral, functional connectivity, and topological descriptors resulted in a high-dimensional feature space containing 1,043 features for each subject. High-dimensional feature spaces often include redundant and irrelevant variables, which may increase computational complexity, **reduce model generalization**, and increase the likelihood of overfitting, particularly when the number of features exceeds the number of subjects [37,38]. Therefore, a feature selection stage was incorporated prior to machine learning classification.

To identify the most informative feature selection strategy, six approaches were comparatively evaluated: Mutual Information, Analysis of Variance (ANOVA), Recursive Feature Elimination (RFE) with Logistic Regression, RFE with Random Forest, RFE with Extra Trees, and RFE with Linear Support Vector Machine (Linear SVM). Each method was evaluated using an identical machine learning pipeline consisting of feature standardization, feature selection, and an RBF Support Vector Machine (SVM) classifier under five-fold stratified cross-validation, ensuring a fair comparison across all feature selection strategies.

Based on the comparative evaluation, Recursive Feature Elimination (RFE) with Logistic Regression achieved the highest overall classification performance and was therefore selected for the proposed framework. RFE is a wrapper-based feature selection algorithm that recursively removes the least informative features according to the importance assigned by the learning algorithm until the desired number of features is retained [39]. Compared with filter-based methods, wrapper-based approaches explicitly consider feature interactions during the selection process, often leading to improved predictive performance.

In the proposed framework, 110 features were retained for all subsequent experiments. The number of retained features was determined empirically through preliminary experiments by comparing different feature subset sizes under the same five-fold cross-validation protocol. A subset of 110 features provided a favorable balance between classification performance and dimensionality reduction and was therefore adopted throughout the remainder of the study. The selected feature subset was subsequently used for all machine learning classifiers, the ablation study, and the Explainable Artificial Intelligence analyses presented in this study.

### 2.7 Machine Learning Classification

The discriminative performance of the extracted EEG features was evaluated using five supervised machine learning classifiers: Support Vector Machine (SVM) [40], Random Forest (RF) [41], Extreme Gradient Boosting (XGBoost) [42], CatBoost [43], and Multi-Layer Perceptron (MLP) [44]. To ensure a fair comparison among classifiers, all models were evaluated using the same feature representation, preprocessing pipeline, feature selection strategy, and validation protocol.

Before classification, the subject-level feature vectors were standardized using the StandardScaler to normalize the feature distributions. Feature selection was subsequently performed using Recursive Feature Elimination (RFE) with Logistic Regression as the base estimator (solver = *liblinear*, maximum iterations = 5000). RFE was configured with a step size of 25 and selected the 110 most informative features, as determined in the feature selection analysis. To prevent information leakage, feature scaling, feature selection, and classifier training were integrated into a single Scikit-learn [45] Pipeline, ensuring that all preprocessing steps were performed independently within each cross-validation fold.

Model performance was assessed using stratified five-fold cross-validation, with sample shuffling enabled and a fixed random seed (random_state = 42) to ensure reproducibility while preserving the class distribution across folds. Classification performance was evaluated using Accuracy, Recall, F1-score, Balanced Accuracy, Receiver Operating Characteristic Area Under the Curve (ROC-AUC) [46], and Precision–Recall Area Under the Curve (PR-AUC) [47]. In addition, Receiver Operating Characteristic (ROC) curves, Precision–Recall (PR) curves, and confusion matrices were generated to provide a comprehensive evaluation of classifier performance.

The evaluated classifiers were configured using the hyperparameters summarized in Table 2.

**Table 2.** Hyperparameter settings of the evaluated machine learning classifiers.

| Classifier | Hyperparameter Settings |
| --- | --- |
| **Support Vector Machine (SVM)** | Kernel = RBF, C = 1, $\gamma$ = scale, probability = True |
| **Random Forest (RF)** | Number of trees = 500, max_depth = None, random_state = 42 |
| **XGBoost** | Number of trees = 300, max_depth = 4, learning_rate = 0.05, subsample = 0.8, colsample_bytree = 0.8 |
| **CatBoost** | Iterations = 300, depth = 6, learning_rate = 0.05 |
| **Multi-Layer Perceptron (MLP)** | Hidden layers = (128, 64), ReLU activation, maximum iterations = 1000 |
| **Feature Selection** | Recursive Feature Elimination (Logistic Regression, liblinear, max_iter = 5000, step = 25, selected features = 110) |
| **Cross-validation** | Stratified five-fold cross-validation, shuffle = True, random_state = 42 |

### 2.8 Ablation Study

To investigate the contribution of each feature domain to the overall classification performance, an ablation study was conducted using the same machine learning framework employed for the final classification model. Ablation analysis is a widely used evaluation strategy that systematically removes or combines feature groups to quantify their individual and collective contributions to model performance [48].

Seven experimental configurations were evaluated: spectral features only, functional connectivity (wPLI) features only, topological (TDA) features only, spectral + wPLI, spectral + TDA, wPLI + TDA, and spectral + wPLI + TDA. For each configuration, the corresponding feature subset was extracted from the subject-level feature matrix before feature selection.

To ensure a fair comparison across all experiments, the same preprocessing pipeline, Recursive Feature Elimination (RFE) feature selection strategy, and Support Vector Machine (SVM) classifier were applied to every feature combination. The number of selected features, cross-validation protocol, and evaluation metrics remained identical to those used in the primary classification experiments. Consequently, differences in classification performance could be attributed solely to the contribution of the included feature domains rather than changes in model configuration or evaluation methodology.

The ablation study enabled the assessment of the individual discriminative power of spectral, functional connectivity, and topological descriptors, as well as the extent to which combining these complementary representations improved subject-level ADHD classification.

### 2.9 Explainable Artificial Intelligence

Although machine learning models can achieve high predictive performance, their decision-making processes are often difficult to interpret, particularly in biomedical applications where transparency and clinical trust are essential. To improve the interpretability of the proposed classification framework, two complementary Explainable Artificial Intelligence (XAI) techniques, SHapley Additive exPlanations (SHAP) and Local Interpretable Model-agnostic Explanations (LIME), were employed to explain the predictions of the final classification model [25,26].

SHAP was used to provide global model interpretation by quantifying the contribution of each selected feature to the overall classification performance. Based on cooperative game theory, SHAP assigns an importance value to every feature according to its contribution to the model prediction, allowing the identification of the most influential EEG biomarkers across the entire dataset [25]. In the present study, SHAP was applied to the final SVM classifier after Recursive Feature Elimination (RFE), enabling the ranking of the 110 selected features according to their average contribution to the classification process.

To complement the global interpretation provided by SHAP, LIME was employed to generate local explanations for individual subjects. LIME approximates the prediction of the trained classifier using an interpretable local surrogate model, thereby identifying the features that contributed most strongly to the prediction of a specific subject [26]. Representative correctly and incorrectly classified ADHD and healthy control subjects were analyzed to investigate the factors influencing both successful and erroneous predictions.

The combined use of SHAP and LIME provides complementary perspectives on model interpretability. While SHAP identifies the global importance of EEG features across the entire dataset, LIME explains individual predictions at the subject level. Together, these approaches improve the transparency of the proposed classification framework and facilitate the identification of neurophysiologically meaningful biomarkers associated with ADHD.

## 3. Results

The proposed framework was evaluated using the subject-level EEG dataset consisting of 121 participants. Feature extraction generated an initial feature space containing 1,043 descriptors, including spectral, functional connectivity, and topological features. Following feature selection, the selected features were used to train and evaluate multiple machine learning classifiers using stratified five-fold cross-validation. The experimental results are presented in the following sections.

### 3.1 Feature Selection Comparison

To identify the most informative subset of EEG features, six feature selection approaches were initially compared, including Mutual Information (MI), Analysis of Variance (ANOVA), Recursive Feature Elimination (RFE) with Logistic Regression, RFE with Random Forest, RFE with Extra Trees, and RFE with Linear Support Vector Machine (Linear SVM). Each method was evaluated using an identical Support Vector Machine (SVM) classifier and stratified five-fold cross-validation protocol to ensure a fair comparison.

Among the evaluated approaches, RFE with Logistic Regression consistently achieved the highest classification performance. Consequently, this method was selected as the feature selection strategy for the remainder of the study. The algorithm reduced the original feature space from 1,043 features to 110 features, substantially decreasing feature dimensionality while preserving the discriminative information required for subject-level ADHD classification.

The selected feature subset served as the input for all subsequent machine learning experiments, ablation analyses, and Explainable Artificial Intelligence investigations.

**Table 3.**
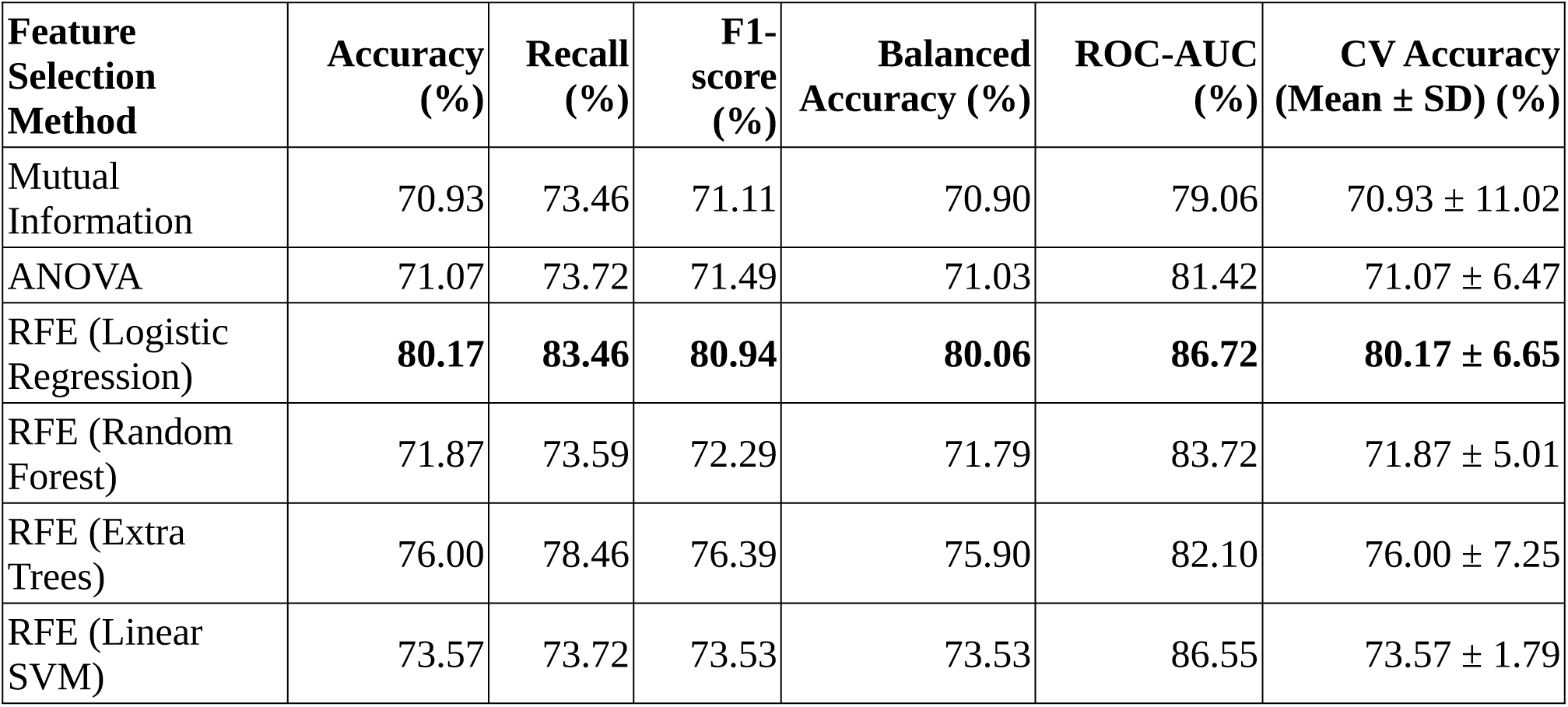
Performance comparison of feature selection methods for subject-level ADHD classification using stratified five-fold cross-validation.

| **Feature Selection Method** | **Accuracy (%)** | **Recall (%)** | **F1-score (%)** | **Balanced Accuracy (%)** | **ROC-AUC (%)** | **CV Accuracy (Mean ± SD) (%)** |
| --- | --- | --- | --- | --- | --- | --- |
| Mutual Information | 70.93 | 73.46 | 71.11 | 70.90 | 79.06 | 70.93 ± 11.02 |
| ANOVA | 71.07 | 73.72 | 71.49 | 71.03 | 81.42 | 71.07 ± 6.47 |
| RFE (Logistic Regression) | **80.17** | **83.46** | **80.94** | **80.06** | **86.72** | **80.17 ± 6.65** |
| RFE (Random Forest) | 71.87 | 73.59 | 72.29 | 71.79 | 83.72 | 71.87 ± 5.01 |
| RFE (Extra Trees) | 76.00 | 78.46 | 76.39 | 75.90 | 82.10 | 76.00 ± 7.25 |
| RFE (Linear SVM) | 73.57 | 73.72 | 73.53 | 73.53 | 86.55 | 73.57 ± 1.79 |

### 3.2 Machine Learning Classification Performance

To evaluate the effectiveness of the proposed EEG feature representation, five supervised machine learning classifiers were trained and tested using the 110 features selected by Recursive Feature Elimination (RFE). The evaluated classifiers included Support Vector Machine (SVM), Random Forest (RF), XGBoost, CatBoost, and Multi-Layer Perceptron (MLP). All models were assessed using the same stratified five-fold cross-validation protocol to ensure a fair comparison.

The classification performance of the evaluated models is summarized in Figure 2. Among all classifiers, the Support Vector Machine (SVM) achieved the highest overall performance, obtaining an accuracy of 80.17%, a recall of 83.61%, an F1-score of 80.95%, and a balanced accuracy of 80.14%. In addition, the SVM achieved a ROC-AUC of 86.72% and a PR-AUC of 87.34%, indicating strong discriminative capability for distinguishing ADHD subjects from healthy controls.

**Figure 2.**
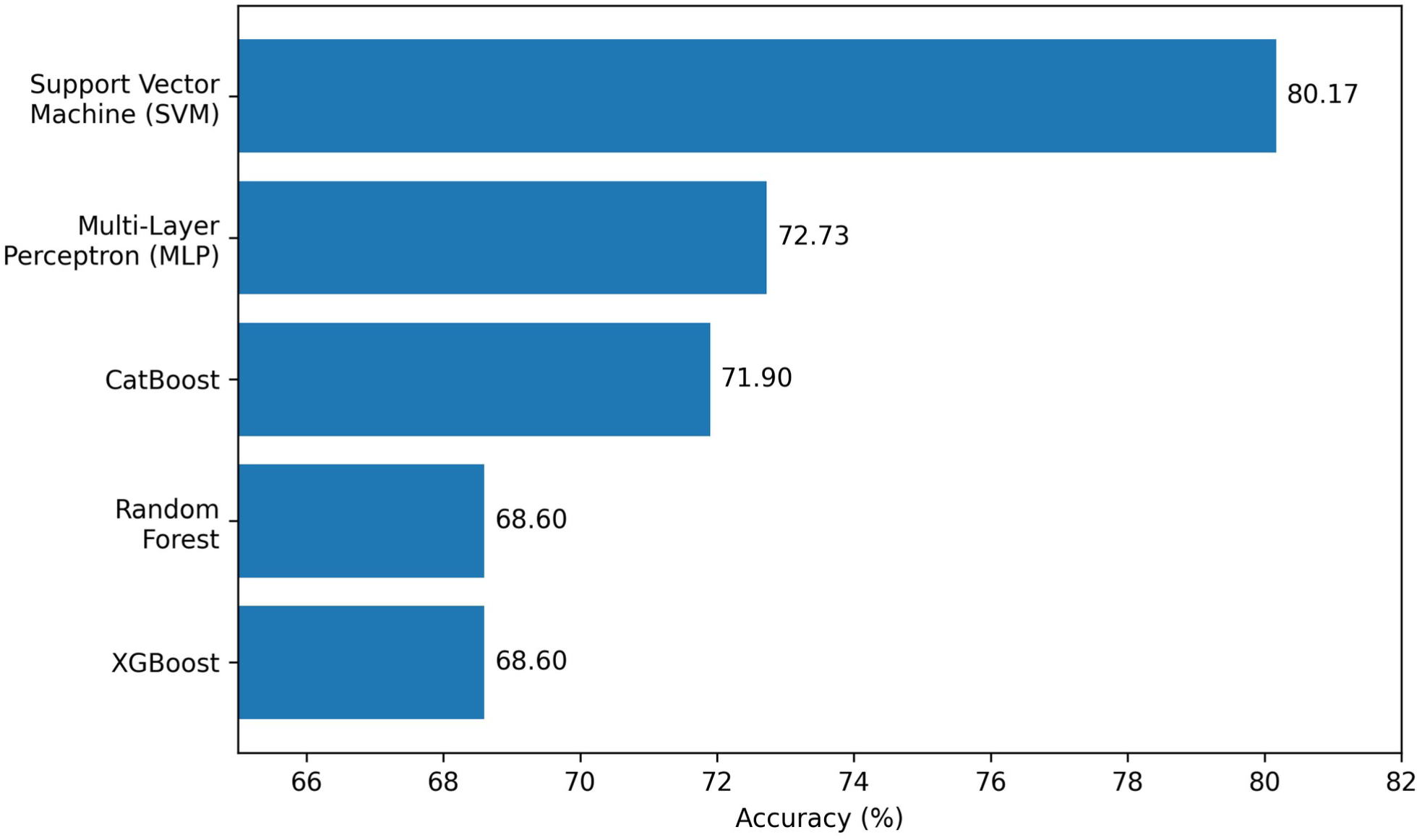
Classification accuracy of the evaluated machine learning classifiers using the selected EEG features under stratified five-fold cross-validation. Among the evaluated models, the Support Vector Machine (SVM) achieved the highest accuracy (80.17%), followed by the Multi-Layer Perceptron (MLP), CatBoost, Random Forest (RF), and XGBoost. The superior performance of the SVM supports its selection as the final classifier for the subsequent ablation study and Explainable Artificial Intelligence analyses.

**Figure 3.**
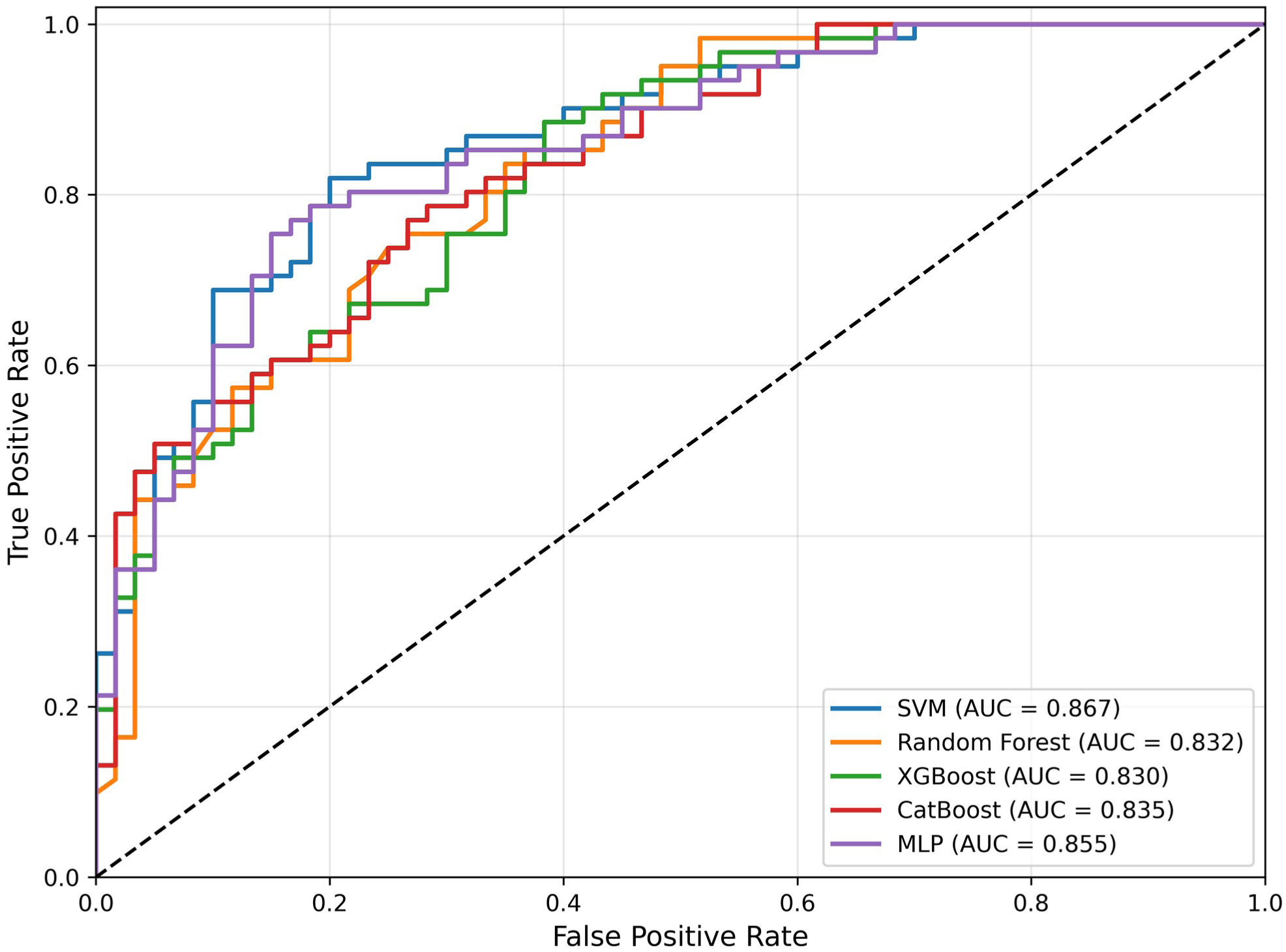
Receiver operating characteristic (ROC) curves of the evaluated machine learning classifiers using the selected EEG features under stratified five-fold cross-validation. The Support Vector Machine (SVM) achieved the highest classification performance (ROC-AUC = 0.867), followed by the Multi-Layer Perceptron (MLP), CatBoost, Random Forest, and XGBoost. The superior ROC-AUC obtained by the SVM indicates its stronger discriminative capability for distinguishing ADHD from healthy control subjects over a wide range of classification thresholds.

Although the remaining classifiers also demonstrated competitive performance, none consistently outperformed the SVM across all evaluation metrics. Consequently, the SVM was selected as the final classification model and was subsequently employed in the ablation study and Explainable Artificial Intelligence analyses.

### 3.3 Ablation Study

To investigate the contribution of each feature category to ADHD classification, an ablation study was conducted using seven experimental configurations: (i) spectral features only, (ii) topological (TDA) features only, (iii) functional connectivity (wPLI) features only, (iv) spectral + TDA, (v) wPLI + TDA, (vi) spectral + wPLI, and (vii) the complete feature set consisting of spectral, wPLI, and TDA features. All experiments employed the same preprocessing pipeline, recursive feature elimination (RFE), Support Vector Machine (SVM) classifier, and stratified five-fold cross-validation to ensure a fair comparison.

**Table 4.** Performance comparison of different feature combinations in the ablation study. The proposed framework integrating spectral, weighted Phase Lag Index (wPLI), and persistent homology (TDA) features achieved the highest overall classification performance, demonstrating that the three feature types provide complementary information for ADHD detection.

| Feature Combination | Accuracy (%) | Recall (%) | F1-score (%) | Balanced Accuracy (%) | ROC-AUC (%) | PR-AUC (%) |
| --- | --- | --- | --- | --- | --- | --- |
| Spectral | 67.77 | 70.49 | 68.80 | 67.72 | 69.52 | 64.02 |
| TDA | 59.50 | 62.30 | 60.40 | 59.47 | 66.16 | 66.31 |
| wPLI | 76.86 | 75.41 | 76.03 | 76.85 | 83.66 | 86.76 |
| Spectral + TDA | 69.42 | 72.13 | 70.97 | 69.39 | 77.07 | 70.85 |
| wPLI + TDA | 75.21 | 75.41 | 75.00 | 75.20 | 84.19 | 86.19 |
| Spectral + wPLI | 78.51 | 78.69 | 78.69 | 78.50 | 85.46 | 87.43 |
| **Spectral + wPLI + TDA (Proposed)** | **80.17** | **83.61** | **80.95** | **80.14** | **86.72** | **87.34** |

### 3.4 Explainability Analysis

To improve the interpretability of the proposed ADHD classification framework, Explainable Artificial Intelligence (XAI) techniques were applied to the best-performing model. The Support Vector Machine (SVM), trained using the optimal feature subset selected by recursive feature elimination (RFE), was analyzed using SHapley Additive exPlanations (SHAP) and Local Interpretable Model-agnostic Explanations (LIME). SHAP was employed to provide a global interpretation by quantifying the contribution of each selected feature across all subjects, whereas LIME was used to generate local explanations for individual predictions. Together, these complementary approaches provide insight into both the overall decision-making process of the classifier and the factors influencing specific classification outcomes.

**Figure 4.**
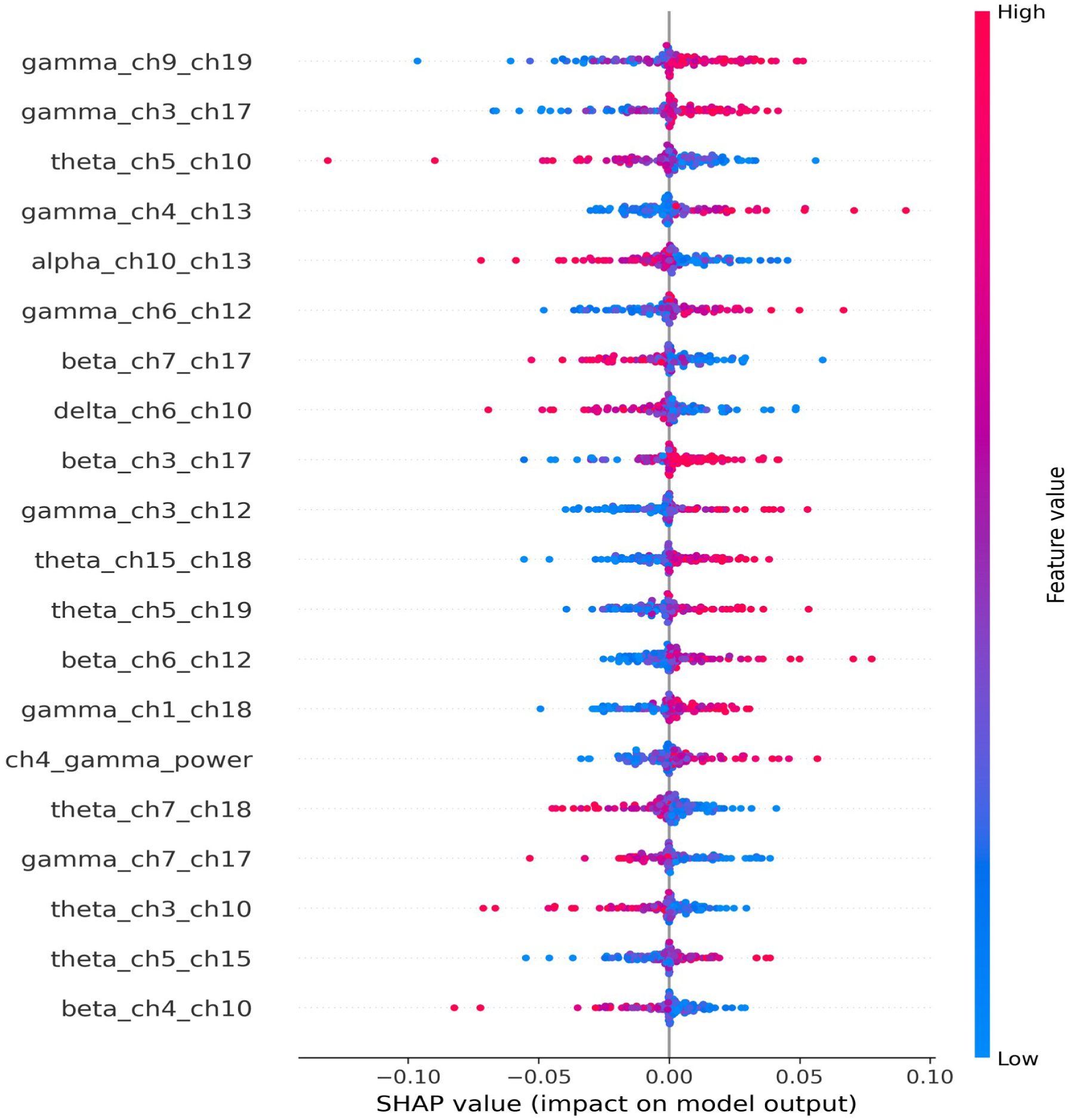
SHAP summary plot illustrating the global importance of the selected EEG features for the proposed Support Vector Machine (SVM) classifier. Features are ranked according to their mean absolute SHAP values. Each point represents an individual subject, with color indicating the normalized feature value (blue: low; red: high). Positive SHAP values increase the likelihood of the ADHD class, whereas negative SHAP values contribute toward the control class.

**Figure 5.**
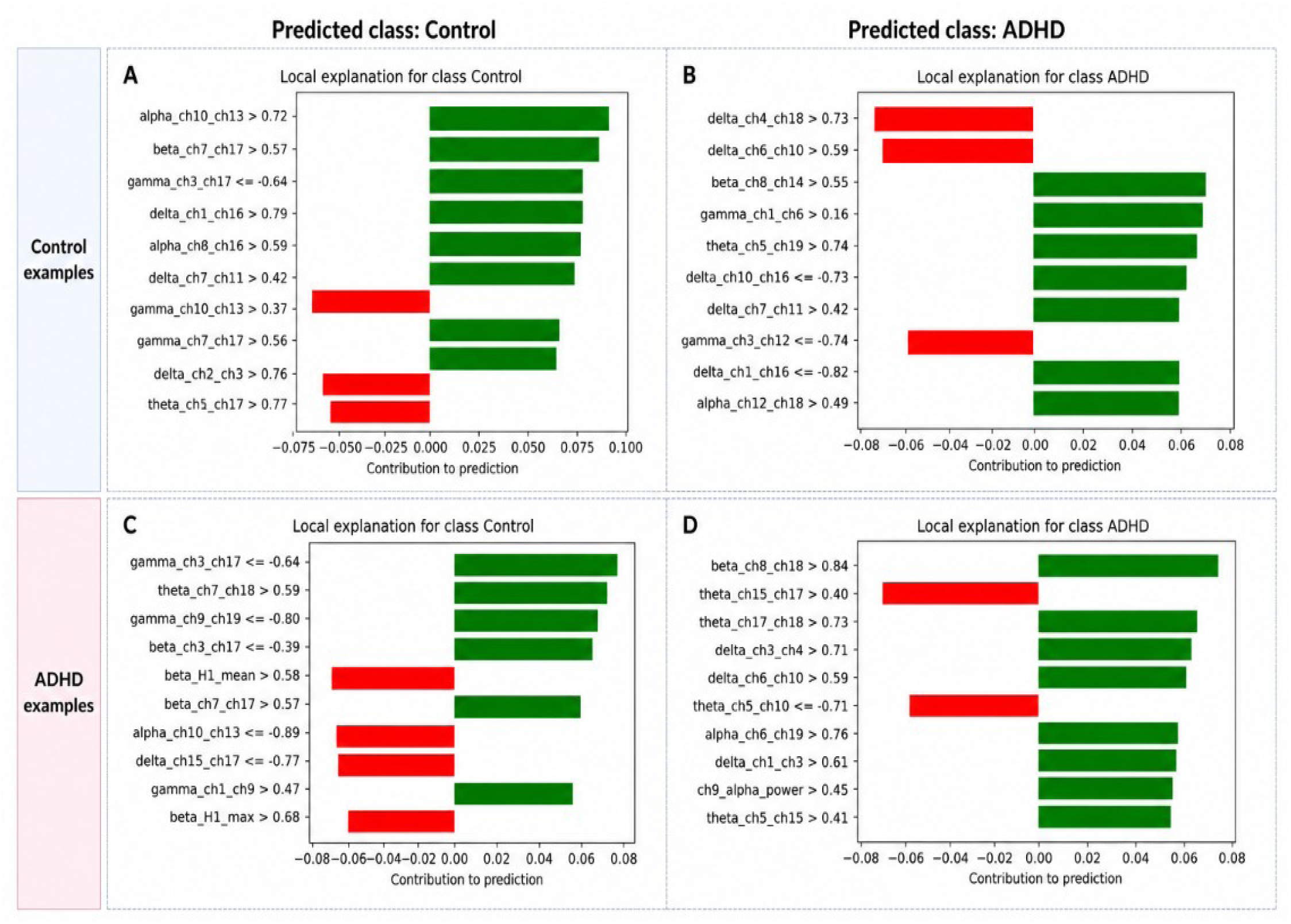
Local Interpretable Model-agnostic Explanations (LIME) for representative EEG subjects classified by the proposed Support Vector Machine (SVM) model. (A, C) Representative control subjects correctly classified as the control class, and (B, D) representative ADHD subjects correctly classified as the ADHD class. Green bars represent features that support the predicted class, whereas red bars represent features that oppose the prediction. The length of each bar indicates the magnitude of the feature’s contribution to the local prediction, demonstrating how spectral, functional connectivity (wPLI), and topological (TDA) features jointly influence individual classification decisions.

## 4. Discussion

The present study introduced a multimodal EEG-based framework for ADHD classification that integrates spectral, functional connectivity, and topological features within a machine learning pipeline. By combining weighted Phase Lag Index (wPLI)-based connectivity analysis with persistent homology-derived topological descriptors and conventional spectral features, the proposed approach achieved an overall classification accuracy of 80.17% and a ROC-AUC of 86.72% using a Support Vector Machine (SVM) classifier. These findings demonstrate that integrating complementary EEG feature representations improves discrimination between ADHD and healthy control subjects.

The ablation study further demonstrated the complementary nature of the extracted features. Although functional connectivity features alone achieved relatively strong performance, combining them with spectral and topological descriptors consistently improved classification accuracy and ROC-AUC. The best results were obtained when all three feature categories were integrated, indicating that each feature type captures distinct aspects of the underlying neural activity associated with ADHD.

An important contribution of this work is the incorporation of Topological Data Analysis (TDA) through persistent homology. Unlike conventional statistical or connectivity-based descriptors, persistent homology characterizes the global geometric organization of EEG-derived connectivity networks across multiple spatial scales. These topological descriptors provide complementary information that is not directly captured by spectral power or pairwise functional connectivity measures, thereby enhancing the robustness of the classification model.

The explainability analysis demonstrated that the proposed framework is not only accurate but also interpretable. SHAP analysis identified the most influential features contributing to the global model behavior, while LIME provided subject-specific explanations for individual predictions. Together, these complementary explainability techniques showed that the classifier relies on meaningful combinations of spectral, connectivity, and topological features rather than isolated biomarkers. Such interpretability is particularly important for clinical decision-support systems, where understanding the rationale behind model predictions can increase clinician confidence and facilitate future clinical translation.

Compared with previous EEG-based ADHD classification studies relying solely on spectral analysis or functional connectivity, the proposed framework provides a more comprehensive representation of brain activity by integrating complementary feature domains. The use of weighted Phase Lag Index reduces the influence of volume conduction on connectivity estimation, while persistent homology captures higher-order network organization that is often overlooked by traditional graph-theoretic approaches. These methodological advances contribute to the improved classification performance observed in this study.

Despite these promising findings, several limitations should be acknowledged. First, the proposed framework was evaluated using a single publicly available EEG dataset, and additional validation on independent cohorts is required to assess its generalizability. Second, the proposed framework was evaluated using EEG recordings acquired during a visual attention task. Future studies should investigate the generalizability of the proposed framework using additional experimental paradigms, including resting-state EEG and other cognitive tasks. Third, although the proposed framework achieved encouraging performance, deep learning architectures trained on substantially larger datasets may further improve classification accuracy.

Furthermore, ADHD is a clinically and neurobiologically heterogeneous disorder, with considerable variability in symptom presentation, developmental trajectory, and underlying neural mechanisms. Consequently, the spectral, functional connectivity, and topological patterns identified in this study should not be interpreted as universal biomarkers applicable to all individuals with ADHD. Rather, they represent discriminative characteristics observed within the analyzed dataset under the proposed experimental framework. Future studies involving larger and more diverse cohorts, external validation datasets, and ADHD subtype-specific analyses are needed to evaluate the robustness, generalizability, and clinical applicability of the proposed framework across heterogeneous patient populations. In addition, future research should investigate multi-center datasets, multimodal neuroimaging integration, and longitudinal analyses to further strengthen the robustness and potential clinical utility of EEG-based ADHD classification.

Overall, the results demonstrate that integrating spectral, functional connectivity, and topological EEG features within an explainable machine learning framework provides an effective and interpretable approach for automated ADHD classification. Although further validation on independent and more diverse datasets is required before clinical deployment, the proposed methodology offers a promising direction for the development of future computer-aided diagnostic systems that combine high predictive performance with transparent and interpretable decision-making.

## 5. Conclusion

This study presented an explainable machine learning framework for automated ADHD classification using EEG signals. The proposed approach integrates spectral features, weighted Phase Lag Index (wPLI)-based functional connectivity, and persistent homology-derived topological descriptors to capture complementary characteristics of brain activity. After recursive feature elimination (RFE) and classification using a Support Vector Machine (SVM), the proposed framework achieved an accuracy of 80.17%, a balanced accuracy of 80.14%, an F1-score of 80.95%, and a ROC-AUC of 86.72%, outperforming the alternative machine learning models evaluated in this study.

The ablation study demonstrated that combining spectral, connectivity, and topological features consistently improved classification performance compared with using any individual feature category or partial feature combinations. Furthermore, SHAP and LIME analyses provided both global and local interpretability of the model, demonstrating that the classifier relies on meaningful combinations of EEG-derived biomarkers rather than isolated features. These explainability analyses increase the transparency of the proposed framework and support its potential use as a clinically interpretable decision-support tool.

Overall, the findings indicate that integrating conventional EEG features with topological data analysis and functional connectivity provides a comprehensive representation of neural activity that improves automated ADHD classification. The proposed framework offers a promising balance between predictive performance and interpretability, making it suitable for future clinical decision-support applications.

Future work will focus on validating the proposed framework using larger multi-center EEG datasets, investigating additional connectivity measures and advanced topological descriptors, and extending the approach through multimodal neuroimaging and deep learning techniques. Such developments may further improve the robustness, generalizability, and clinical applicability of Explainable Artificial Intelligence systems for ADHD diagnosis.

## Declarations

### Ethics statement

This study used only openly available, fully de-identified human data that were originally located at https://doi.org/10.21227/rzfh-zn36 (IEEE DataPort, “EEG data for ADHD / Control children”, Nasrabadi et al., 2020) [28]. No new data were collected and the authors had no access to identifying information; therefore, no additional ethics approval was required for this secondary analysis.

### Data availability

The EEG data analysed in this study are openly available on IEEE DataPort (“EEG data for ADHD / Control children”, Nasrabadi et al., 2020) at https://doi.org/10.21227/rzfh-zn36 [28].

### Code availability

The analysis code is available from the corresponding author upon reasonable request.

### Author contributions

F.R.: Conceptualization, Methodology (machine learning and explainable AI), Writing – original draft, Writing – review & editing. S.M.: Software, Formal analysis, Writing – original draft. P.E.: Software, Writing – original draft. R.Z.: Validation (neuroscientific interpretation), Writing – original draft, Writing – review & editing. A.M.: Validation (neuroscientific interpretation), Writing – original draft, Writing – review & editing. All authors read and approved the final manuscript.

## Acknowledgments

The authors thank Nasrabadi et al. for making the ADHD/control EEG dataset openly available.

